# fMRI as an outcome measure in clinical trials: A systematic review in clinicaltrials.gov

**DOI:** 10.1101/19002972

**Authors:** Alaleh Sadraee, Martin P. Paulus, Hamed Ekhtiari

## Abstract

**Background:** Functional magnetic resonance imaging (fMRI) is quickly becoming a significant outcome measure for clinical trials and as more than one thousand trials with fMRI as an outcome measure were registered in clinicaltrials.gov at the time of writing this article. However, 93% of these registered trials are still not completed with published results and there is no picture available about methodological dimensions of these ongoing trials with fMRI as an outcome measure.

**Methods:** We collected trials that use fMRI as an outcome measure by searching “fMRI” in the ClinicalTrials.gov registry on October 13 2018 and reviewing each trial’s record entry. Eligible trials’ characteristics were extracted and summarized.

**Results:** In total, 1386 clinical trials were identified that reported fMRI in their outcome measures with fMRI as the only primary outcome in 33% of them. 82% of fMRI trials were started after 2011. The most frequent intervention was drug (29%). 57% of trials had parallel assignment design and 20% were designed for cross over assignment. For task-based fMRI, cognitive systems (46%) based on RDoC was the most frequent domain of tasks, followed by positive valence systems (19%), systems for social processing (10%) and sensorimotor systems (5%). Less than one-third of trials (28%) registered at least one region of interest for their analysis. Food cue reactivity task, pain perception task, n-back task and monetary incentive delay task were recruited in more than 25 registered trials.

**Conclusion:** The number of fMRI trials (fMRI as an outcome measure) with both task and rest protocols is growing rapidly. Different RDoC domains are covered by various tasks in fMRI trials. However, our study suggests the need of greater harmony and better standardization for registration of fMRI details on both methods and analysis which would allow for more effective comparison across studies in systematic reviews and also help the validation of results towards having fMRI as a biomarker in the future.

## Introduction

FMRI is the most powerful and dominant imaging technique in living human brain by now that have entered a variety of branches of today’s clinical research to apply new advances in clinical practice. Speaking of clinical trials, fMRI’s involvement in different domains is so acknowledged that in the current NIH case studies regarding their 2014 definition of clinical trials, there are specific case studies for fMRI in 6 variations trying to elucidate the distinction of the different roles of fMRI in clinical trials as a clinical measurement tool and also as an intervention (NIH, 2018, case #18” a-f”).

In our expectation, the role of fMRI in clinical trials would hopefully be as a biomarker by definition, that is, “a characteristic that is objectively measured and evaluated as an indicator of normal biological processes, pathogenic processes, or pharmacologic responses to a therapeutic intervention” (Biomarkers Definitions Working Group, 2001). To be more specific, fMRI is supposed to be used in order to facilitate development of new interventions by objectively monitoring functional changes in brain, hypothetically targeted by the interventions. For example, changes in specific brain activity after taking pharmacological agents are detected by fMRI to evaluate the effect of treatment in early phases in drug trials. In this regard, fMRI would be promising for accelerating the development of new treatment; In drug development for mental health disorders, urgent need for new treatment was address by an NIMH program, Fast-Fail trials (FAST), utilizing target-engagement biomarkers including fMRI as one of the potential functional biomarker (https://www.nimh.nih.gov/research-priorities/research-initiatives/fast-fast-fail-trials.shtml). FMRI also might play an exploratory role in clinical trials to providing more objective evidence base for submission to regulators, e.g. in later phases of drug trials (Carmichael et al., 2017). In this study we examined fMRI as an outcome measure, but speaking more broadly, by NIH definition of intervention—manipulation of the subject or subject’s environment for the purpose of modifying one or more health-related biomedical or behavioral processes and/or endpoints—it seems that fMRI can also be considered as an intervention in clinical trials in certain conditions (e.g. pre-surgical mapping) (NIH, 2018, case #18e).

Nonetheless, for fMRI to become a powerful biomarker in clinical trials, still some technical and logistical measurements need to be taken in order to make the results reliable and valid. There is a high range of false positive results in fMRI activations (Wager et al., 2009) and the vast choice of analysis are one of the main reasons (Poldrack et al., 2017); Carp (2012) showed that 6,912 unique analysis pipelines are possible for one single dataset which leads to 34,560 significance maps that varies in activation strength, location, and extent. Registration of fMRI methodology before the study or pre-registration which restricts “research’s degree of freedom”—a term coined by Simon (2011), meaning the flexibility of researcher at methodological level— is one of suggested logistical measurements to deal with false positive results (Button et al., 2013; Munafò et al., 2017; Poldrack et al., 2017; Carmichael et al., 2017). Clinical trials’ protocol registration in publicly available clinical trial registries is mandatory by laws and policies (https://clinicaltrials.gov/ct2/about-site/history). ClinicalTrials.gov is the largest clinical trials registry with over 200,000 registered studies. The registry was first established to make possible the public access to clinical trials for participation purposes, and as the need for transparency and reproducibility in science emerges, it extended its goals to also serve as a registry for better monitoring and tracking of clinical trials (Zarin & Keselman, 2007). As the time passed, the site tightened its rules and modified the protocol registration’s required information for more accurate and complete registration. However, for fMRI in clinical trials, the best practices for pre-registration needs to be well-defined through a detailed checklist. If following that checklist becomes obligatory by policy makers and granting agencies as a module in the protocol registration in formal clinical trial registries, like clinicaltrials.gov, hopefully it will result in more valid and replicable fMRI outcomes in clinical trials in long term.

In this study, we completed a systematic review to investigate the scope of fMRI clinical trials in clinicaltrials.gov. We extracted the available data for characteristics of fMRI clinical trials. For categorizing the large number of tasks in task-based fMRI trials, we chose NIMH Research Domain Criteria (RDoC) construct definition (https://www.nimh.nih.gov/research-priorities/rdoc/index.shtml). As a potential pre-registration role that clinicaltrials.gov could play in future for an fMRI specific data element, we also explored the extent of available pre-registered data in clinicaltrials.gov and provided few recommendations.

## Method

### Search strategy and study selection

We searched clinicaltirals.gov on 10/13/2018 for potential clinical trials that used task-based or resting-state fMRI as their outcome measure by using the search term fMRI. Clinicaltrials.gov is a registry that includes more than clinical trials (or interventional studies). Hence, the search was then restricted to clinical trials by applying one of the potential search filters “study type: Interventional”. For further analysis, we downloaded the results in a tab-separated values format and transferred it to an excel table to fix the dataset for later analysis. The dataset encompassed eligible trials’ protocol mandatory required information (data elements) which was submitted to the clinicaltrials.gov registry by sponsor or principle investigator at the start of the study. In order to include completed or ongoing clinical trials we excluded Suspended, Terminated, Withdrawn, and Unknown study states. We also excluded the following studies by manually reviewing each trial’s record page:

- Trials that the outcome measures were recording data of real-time neuro-feedback fMRI to reduce methodological complexities
- Trials with empty (not recorded) outcome measures
- Trials that didn’t report fMRI in their outcome measures either as a consequence of incomplete reporting, or in other cases e.g. if fMRI was used as a part of an intervention in the trial, like “brain surgery based on pre-surgical planning with fMRI”
- Explicitly stated Not BOLD fMRI trials e.g. perfusion fMRI
- Non-relevant trials that appeared in our result as a result of similarity of keywords

Trials’ selection process is outlined in the flow diagram (Figure 1). To follow the best practices in reporting, we applied relevant ‘Preferred Reporting Items for Systematic Reviews and Meta-analyses’ PRISMA guidelines. See Checklist S1 for PRISMA checklist of items.

**Figure 1.**
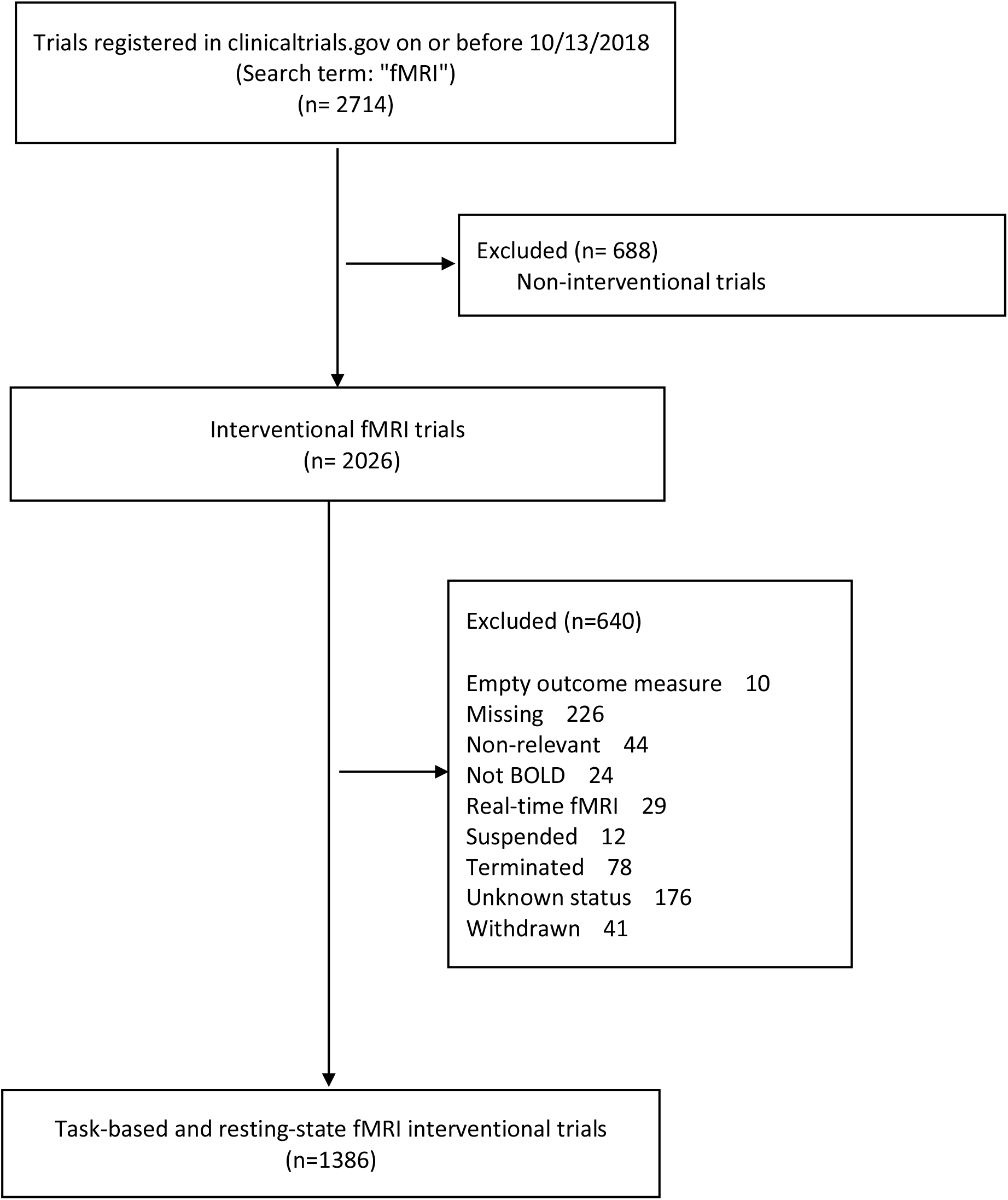
Inclusion flowchart of fMRI trials registered in clinicaltrials.gov on or before 10/13/2018

### Data collection

The following variables are the general characteristic that were available in the dataset:

Recruitment, Start Date, Primary Completion Duration (Year), Location, Funding Source, Gender, Age, Intervention, Primary Purpose, Phase, Intervention Model, Masking, Allocation, Enrollment.

The authors then manually reviewed each trial’s record page for extracting fMRI variables:

FMRI as an Outcome Measure, FMRI Type, Task Name, and Reported Region of Interests.

### Definition and classification of variables

Details of all data elements’ definitions in the XML file are available at the ClinicalTrials.gov (https://prsinfo.clinicaltrials.gov/definitions.html).

#### FMRI as an outcome measure

It defines whether fMRI serves as the “only primary outcome measure” (for clinical trials that have no other primary outcome measures except for fMRI), “one of the primary outcome measures”, “one of the secondary outcome measures” (but not in the primary outcome measures), or “other outcome measures” (but not in the primary or secondary outcome measures) of the trial. It is better to mention that primary outcome measures’ groups also encompass trials that use fMRI as secondary in addition to primary outcome measures, and we didn’t categorize trials which use fMRI in both primary and secondary outcome measures in an individual group due to the importance of primary compared to secondary outcome

#### FMRI method

FMRI method used as an outcome measure is categorized on three main groups of “Resting-state fMRI”, “Task-based fMRI”, “Resting-state fMRI-Task-based fMRI”; Studies that didn’t mention their fMRI method explicitly were labeled as “Not specified” with two exceptions: If the study mentioned the stimuli (e.g. brain response to visual food cues) or an action (e.g. Moving hand, Stimulation) we included them in task-based fMRI.

#### Task name

In order to be able to categorize the tasks, the task name provided by authors’ manipulation (e.g. “brain response to visual food cues” were considered as “Food cue reactivity task”). We categorized tasks within the framework of RDoC. For the sake of clarity, “cognitive systems” domain is broken into its sub-constructs (attention, perception, working memory, declarative memory, language, and cognitive control). Because of the ambiguity in names of the tasks, the ones related to each RDoC domain are classified into three groups of “Well specified”, “Partially specified”, and “Not specified (Table S2).

#### Reported region of interests

We recorded data on whether trials reported at least one major region in which the activation will be examined or have a hypothesis included the activation of at least one region; for resting state fMRI we also included common networks (e.g. DMN) as reported regions of interest. As this section was a part of a bigger plan to examine trials from pre-registration aspect (see the introduction and discussion), we excluded studies with results for this part.

### Statistical analysis

All characteristic data is summarized and analyzed using Excel 2016. Categorical variables are reported as frequencies and percentages.

## Result

The ClinicalTrials.gov registry includes 2026 entries for interventional studies with fMRI as a keyword. Among these, we identified 1386 completed (46%) or ongoing clinical trials (54%) that used BOLD fMRI as an outcome measure. The reasons for exclusion of 640 interventional trials are listed in Figure 1. Among completed trials 84% didn’t report their results yet. In 33% of trials fMRI was the only primary outcome (Table 1). 50% of trials had less than 50 participants and 6% more than 200. 82% of fMRI trials were started after 2011. The most frequent intervention was drug (29%). 57% trials had parallel assignment design and 20% were designed for cross over assignment (Table 2). More details on the characteristics of trials can be found in Table 1 and Table 2.

**Table 1.**
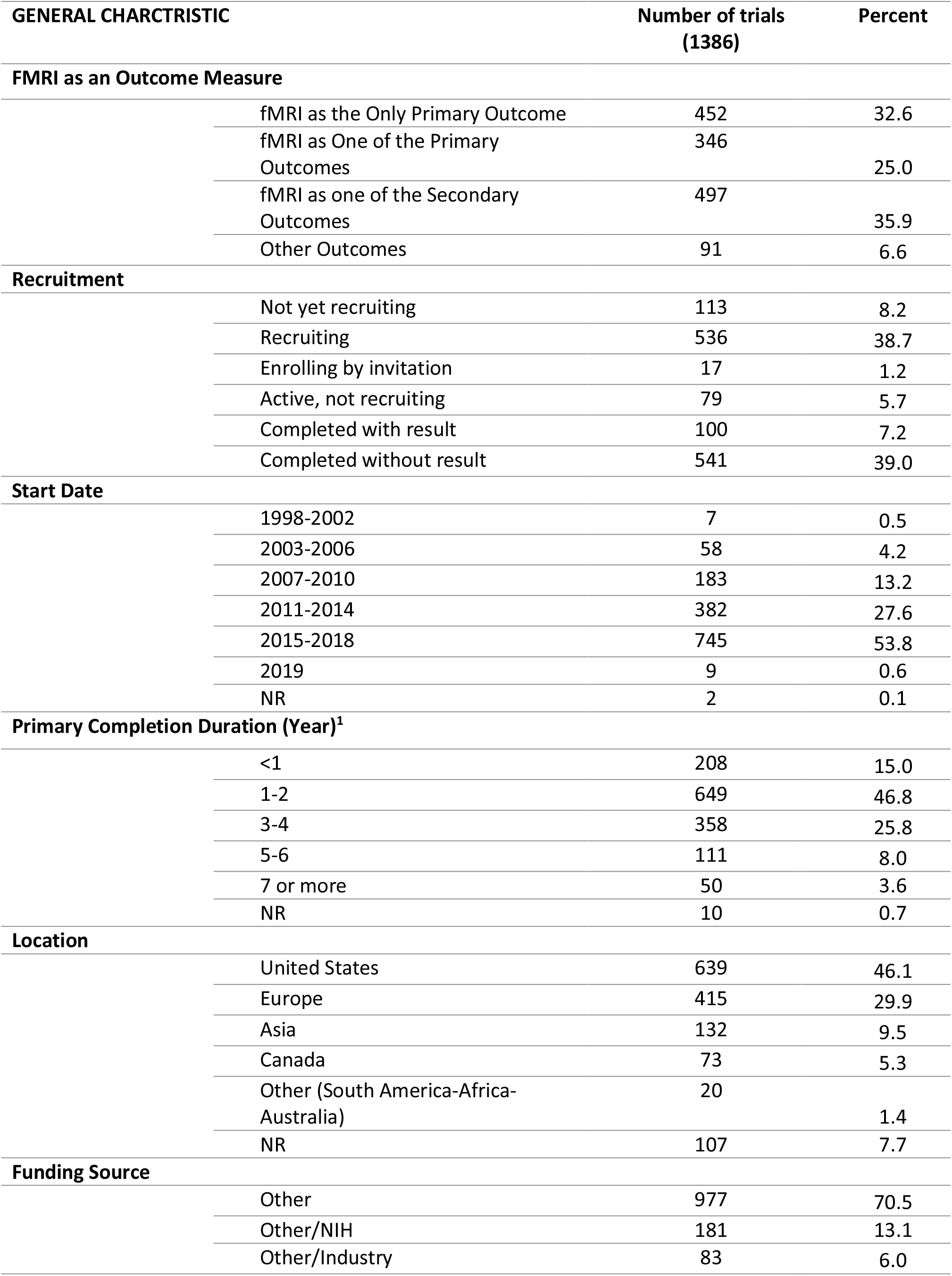
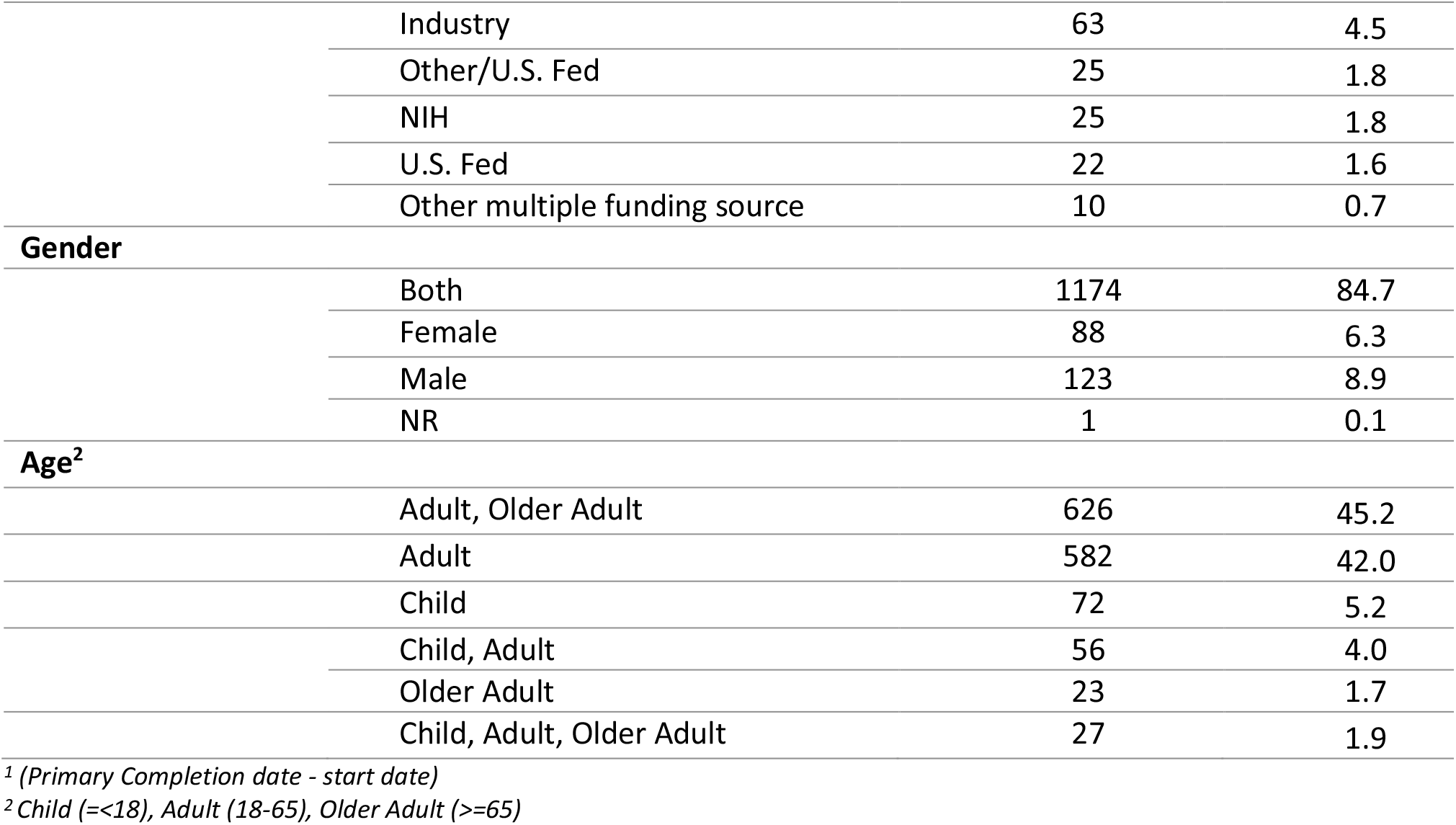
General characteristics of fMRI trials registered in clinicaltrials.gov on or before 10/13/2018

**Table 2.**
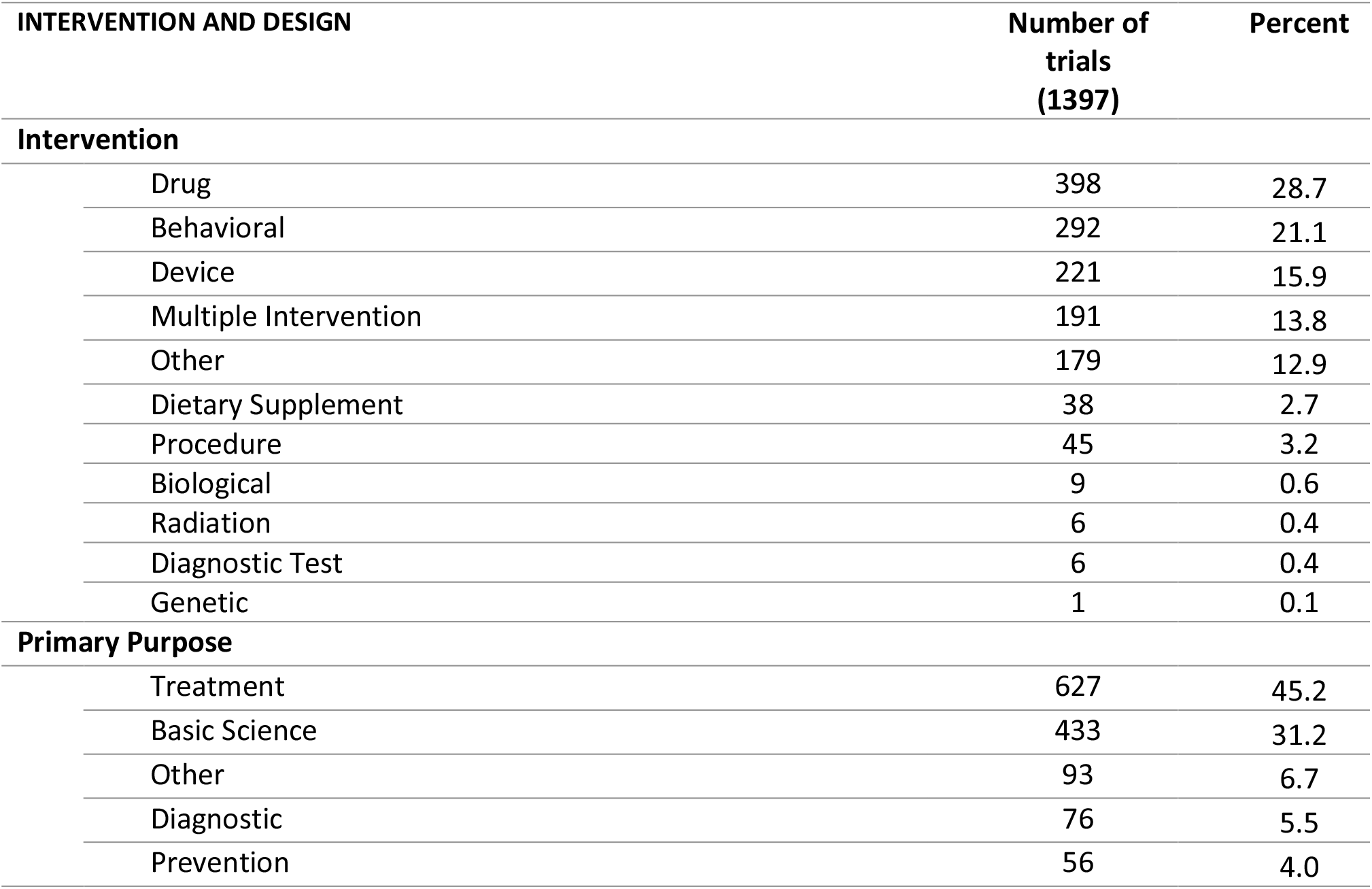
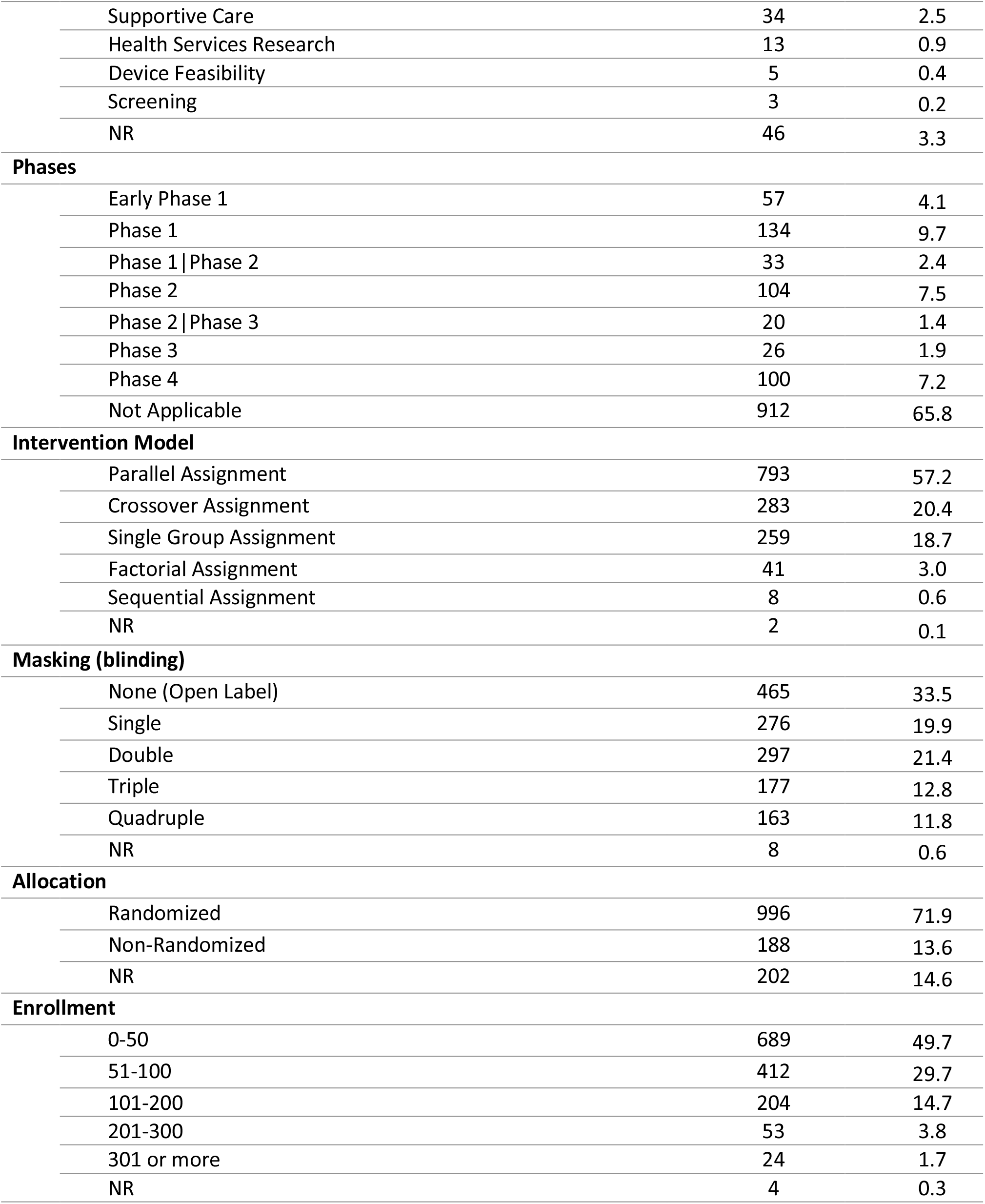
Intervention and design Characteristics of fMRI trials registered in clinicaltrials.gov on or before 10/13/2018

Forty-two percent of trials registered task-based fMRI, 19% resting state, and 12% of trials registered both as the outcome measures. There were 963 tasks in 753 trials that reported task-based fMRI as a part of their outcome measures. 20% of trials used more than one task (Table S1 provides details of the number of tasks in these trials). Positive valence systems (19%), cognitive control (15%), perception (10%), and systems for social processes (10%) were the most frequent domains/subdomains of tasks based on RDoC (Table 3). Food cue reactivity task, pain perception task, n-back task and monetary incentive delay task were recruited in more than 25 registered trials (Table 4). You can find more details on the fMRI tasks in each domain in Table S2. The trend of use in the most frequently recruited fMRI tasks is also reported in Table 5. We found that 28% of trials have reported at least one region of interest. We also included some of the best pre-registered data samples of pre-processing and analysis plan that was found in our clinical trials data-set in Table S3. The dataset is also shared in the Supplementary Material (Supplementary_DataSet).

**Table 3.** RDoC domain of task-based fMRI trials register in clinicaltrials.gov on or before 10/13/201

| RDoC Domain |  | Number of Tasks<br>(963) |  | Percent |
| --- | --- | --- | --- | --- |
| Negative valence systems |  | 50 |  | 5.2 |
| Positive valence systems |  | 186s |  | 19.2 |
| Negative and/or positive valence systems |  | 38 |  | 4.0 |
| Cognitive Systems | Attention | 32 | 3.3 | 45.6 |
|  | Perception | 98 | 10.2 |  |
|  | Declarative memory | 61 | 6.3 |  |
|  | Language | 23 | 2.4 |  |
|  | Cognitive control | 143 | 14.8 |  |
|  | Working memory | 82 | 8.5 |  |
| Systems for social processes |  | 99 |  | 10.3 |
| Arousal/Regulatory Systems |  | 1 |  | 0.1 |
| Sensorimotor Systems |  | 52 |  | 5.4 |
| Not specified |  | 98 |  | 10.2 |

**Table 4.**
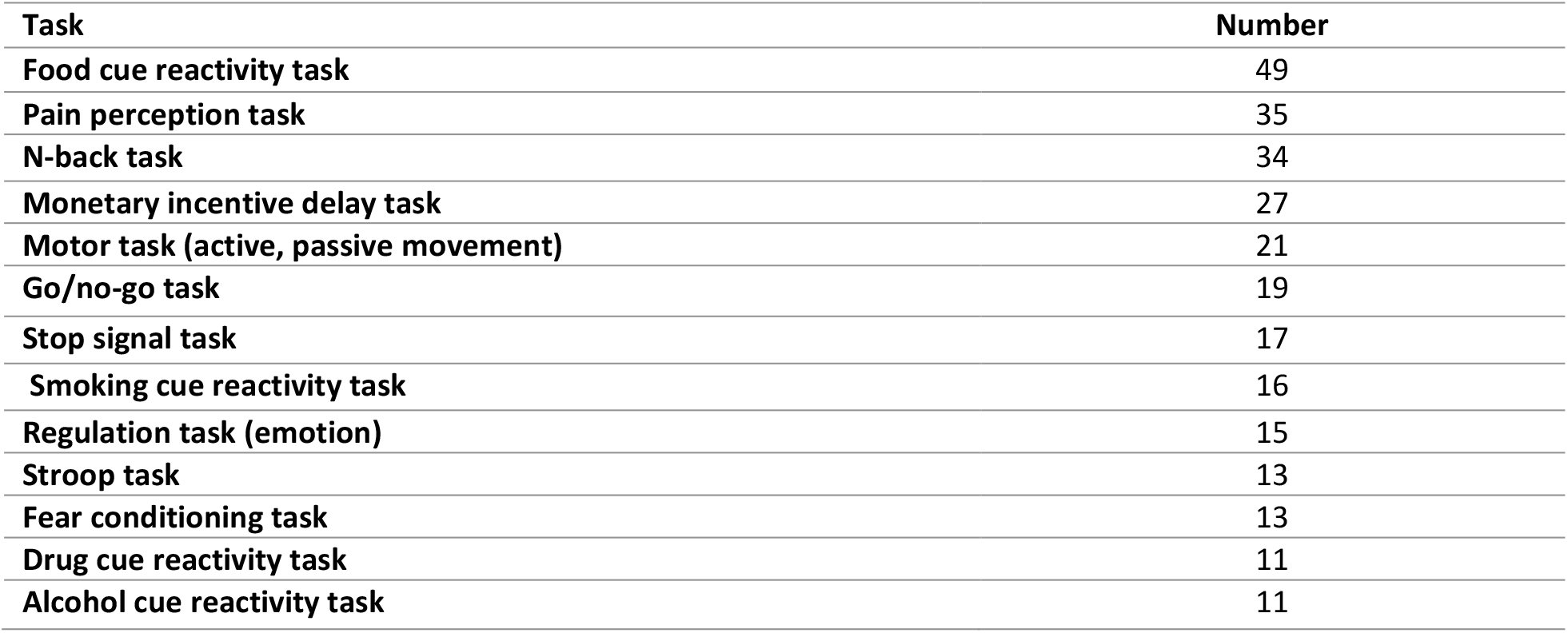
Task classification of task-based fMRI trials registered in clinicaltrials.gov on or before 10/13/2018 based on RDoC domains for well-specified tasks with more than 10 repetitions

**Table 5.** Trend of well-specified tasks with more than 10 repetitions from 2001-2018

| Task | Year | 2001-2003 | 2004-2006 | 2007-2009 | 2010-2012 | 2013-2015 | 2016-2018 |
| --- | --- | --- | --- | --- | --- | --- | --- |
| Cue reactivity task (food) (48)* |  | 0 | 2 | 5 | 12 | 12 | 17 |
| Pain perception task (35) |  | 0 | 4 | 4 | 9 | 7 | 11 |
| N-back task (34) |  | 0 | 2 | 3 | 7 | 10 | 12 |
| Monetary incentive delay (MID) task (27) |  | 0 | 0 | 0 | 5 | 13 | 9 |
| Motor task (active, passive movement) (21) |  | 0 | 2 | 2 | 3 | 6 | 8 |
| Go/no-go task (19) |  | 0 | 0 | 2 | 4 | 4 | 9 |
| Cue reactivity task (smoking) (16) |  | 0 | 1 | 3 | 4 | 4 | 4 |
| Regulation task (emotion) (15) |  | 1 | 0 | 1 | 3 | 5 | 5 |
| Stroop task (13) |  | 0 | 0 | 4 | 5 | 1 | 3 |
| Fear conditioning task (13) |  | 0 | 0 | 0 | 3 | 3 | 7 |
| Cue reactivity task (drug) (11) |  | 0 | 1 | 0 | 3 | 2 | 5 |
| Cue reactivity task (alcohol) (11) |  | 0 | 1 | 1 | 1 | 1 | 7 |
| Task-based fMRI (962)* |  | 5 | 33 | 104 | 150 | 280 | 390 |

One of the studies' start date is missed

## Discussion

In order to develop an understanding of fMRI usage in clinical trials as an outcome measure, we systematically reviewed the fundamental characteristics of eligible trials registered in the ClinicalTrials.gov before 2019. During the last 20 years, fMRI as an outcome measure have grown rapidly such that about half of the total trials with fMRI as an outcome measure were registered after the year 2016. 54% of fMRI trials are still ongoing and 84 % of the completed trials didn’t report their result yet so we still do not have a good methodological representation of these ongoing or completed trials in the published literature. This is the first report to give an overview about the current status in the field based on the available data in clinicaltrials.gov. In about one third of the included trials in this systematic review, fMRI was the only primary outcome measure and the “Primary Purpose” of half of the trials was reported as “treatment”. 29% of trials examined a drug intervention. 70% of trials had randomized allocation. Triple and Quadruple blindness overall made up 25 percent of trials. Cognitive systems (46%) based on RDoC was the most frequent domain of tasks, followed by positive valence systems (19%), systems for social processing (10%), and sensorimotor systems (5%). Emotional processing tasks, cue reactivity tasks, pain perception tasks, N-back task, emotional faces processing task, motor tasks, and Go/no-go task was the most frequent tasks that used in these trials (please see supplementary material 2 for more details). fMRI statistical analysis details in registered trials is scarce and less than one third of trials registered at least one region of interest for their analysis.

We reviewed all registered materials for each trial to extract fMRI specific information that might be reported in different sections such as outcome measures, study arms, detailed description. In about 25 percent of trials the rest or task-based fMRI types were not specified. In about 10 percent of task-based fMRI trials there was not any clear indication (even of task-domain). Task’s name as well as reporting model was varied according to the following categories: 1) Conventional name mentioned explicitly (e.g., monetary incentive delay task) 2) Task was introduced in a general way (e.g., decision-making task, cognitive task) 3) The procedure was indicated (e.g., listen to baby’s cry, exposure to auditory and visual food cues, luminous stimulation) 4) Task’s name was not mentioned but the task’s details was described 5) evaluation of brain response to stimuli in study implicated the task (e.g. brain activity in response to noxious stimuli, as assessed by fMRI). We organized the data to be consistent and placed them in tabular format by categorizing the task domains according to the RDoC definition.

In an effort to search for pre-registered data of fMRI specification, we had a plan to collect details of registered fMRI information including data design, data acquisition, data pre-processing, and analysis plan, but the minute amount of available data and the lack of similarity in reporting style posted restrictions on this procedure so that we were unable to take our pre-registration classification any further (See Table S3). Even for basic details like regions of interest (ROI), less than one third of trials registered at least one ROI for their analysis.

At this point there is sufficient evidence that higher standards of pre-registration information are needed to help transparency and reproducibility of research, which would lead to bias reduction. Overall, as Munafo et al. (2017) have mentioned, study design, primary outcome(s), and analysis plans are what should be pre-specified for a study in the strongest form of pre-registration. There are also some previous efforts to recommend essentials for an fMRI study pre-registration; editors of “Neuroimaging: Clinical” referred to the existence possibility of prediction in clinical neuroimaging studies and pre-registration of ROIs as a specific suggestion against SHARKing. “This is common in clinical trials and there is no reason that strong predictions cannot be defined in clinical neuroimaging studies. Pre-registration of ROIs may be particularly useful as it guards against SHARKing “(Roiser et al., 2016). Poldrack and colleagues (2017) have indicated “planned sample size, specific analysis tools to be used, specification of predicted outcomes, and definition of any specific ROIs or localizer strategies that will be used for analysis” as the details of an fMRI study that should be pre-registered. Particularly in clinical trials —to extend Carmichael et al., (2017) statement is helpful (even though it is initially recommended for drug development trials): “fMRI methodology should be held to the same standard as other clinical endpoints, namely, methods must be pre-specified and fixed for the duration of the study. This pre-specification should include a thorough description of task design and implementation, image acquisition and quality control, data preprocessing, ROI definition, model estimation, and endpoint calculation (Schwarz et al. 2011a, b)”.

The current study is the first one that tried to provide an overview about registered trials with fMRI as an outcome measure in clinicaltrials.gov to the best of our knowledge. It should be mentioned that ClinicalTrials.gov is not the sole registry for all clinical trials in the world, so our database is a limited subset of all fMRI clinical trials. The use of fMRI as an outcome measures in clinical trials is growing fast, imposing the requirement for a considerable amount of registration standardization for fMRI specific information to make the accumulating data useful for researchers and assure the validation of results. Based on this systematic review, we suggest following actions:

### Recommendation 1

fMRI type (task-based fMRI vs resting-state fMRI, and basics of pulse sequence like BOLD EPI) and the detailed description of the task/rest should be provided in a clear and replicable way. Sometimes, even with the label of famous fMRI tasks, investigators recruit various versions with major or minor differences. A consensus on a list of major fMRI tasks with their available codes/stimuli could reduce this variability.

### Recommendation 2

Providing official reference checklist of specific required items for pre-registered clinical trial with fMRI as an outcome measure will be important. Committee on Best Practices in Data Analysis and Sharing (COBIDAS) offered a comprehensive reporting checklist for an fMRI study (Nichols et al., 2017). From authors’ perspective, for a strong form of pre-registration, the content of conventional method section of a reported fMRI study should be specified beforehand (registered-report). Hence, we recommend specifying items in Tables D.1 - D.5 of the COBIDAS reporting checklist for pre-registration.

### Recommendation 3

Preregistration of fMRI analysis details in a replicable way is a labor demanding job. Having a citable list of most acceptable fMRI analysis pipelines within major analysis platforms without wiggling room for further analytics explorations could be very helpful to increase transparency and replicability.

### Recommendation 4

Definition of minimum requirements for fMRI outcome preregistration in major platforms like clinicaltrials.gov and then suggestions of the optimum list of items to be registered is required. Expectation for a comprehensive pre-registration for fMRI analysis in the first step might be significantly complex for investigators with less experience and can act as a serious barrier.

### Recommendation 5

All preregistration efforts should not suppress potentials for innovations in new task design and more efficient fMRI analysis pipeline development. We also support secondary data exploration when it is transparently reported.

Currently, there is a large gap on harmonized collective efforts with shared fMRI protocols and study designs to help accumulate replicable knowledge in the field over the time. We hope this systematic review and its recommendations will help to move one step forward for this endeavor.

## Data Availability

The data-set is available in supplementary materials.

## Acknowledgement

Authors would like to thank Dr. GholamAli Hossein_Zadeh, Iranian National Brain Mapping Laboratory, for his valuable comments on this project.

